# Safety and immunogenicity of a live recombinant Newcastle disease virus-based COVID-19 vaccine (Patria) administered via the intramuscular or intranasal route: Interim results of a non-randomized open label phase I trial in Mexico

**DOI:** 10.1101/2022.02.08.22270676

**Authors:** Samuel Ponce-de-León, Martha Torres, Luis Enrique Soto-Ramírez, Juan José Calva, Patricio Santillán-Doherty, Dora Eugenia Carranza-Salazar, Juan Manuel Carreño, Claudia Carranza, Esmeralda Juárez, Laura E. Carreto-Binaghi, Luis Ramírez-Martínez, Georgina Paz-De la Rosa, Rosalía Vigueras-Moreno, Alejandro Ortiz-Stern, Yolanda López-Vidal, Alejandro E. Macías, Jesús Torres-Flores, Oscar Rojas-Martínez, Alejandro Suárez-Martínez, Gustavo Peralta-Sánchez, Hisaaki Kawabata, Irene González-Domínguez, José Luis Martínez-Guevara, Weina Sun, David Sarfati-Mizrahi, Ernesto Soto-Priante, Héctor Elías Chagoya-Cortés, Constantino López-Macías, Felipa Castro-Peralta, Peter Palese, Adolfo García-Sastre, Florian Krammer, Bernardo Lozano-Dubernard

## Abstract

There is still a need for safe, efficient and low-cost coronavirus disease 2019 (COVID-19) vaccines that can stop transmission of severe acute respiratory syndrome coronavirus 2 (SARS-CoV-2). Here we evaluated a vaccine candidate based on a live recombinant Newcastle disease virus (NDV) that expresses a stable version of the spike protein in infected cells as well as on the surface of the viral particle (AVX/COVID-12-HEXAPRO, also known as NDV-HXP-S). This vaccine candidate can be grown in embryonated eggs at low cost similar to influenza virus vaccines and it can also be administered intranasally, potentially to induce mucosal immunity. We evaluated this vaccine candidate in prime-boost regimens via intramuscular, intranasal, or intranasal followed by intramuscular routes in an open label non-randomized non-placebo-controlled phase I clinical trial in Mexico in 91 volunteers. The primary objective of the trial was to assess vaccine safety and the secondary objective was to determine the immunogenicity of the different vaccine regimens. In the interim analysis reported here, the vaccine was found to be safe and the higher doses tested were found to be immunogenic when given intramuscularly or intranasally followed by intramuscular administration, providing the basis for further clinical development of the vaccine candidate. The study is registered under ClinicalTrials.gov identifier NCT04871737. Funding was provided by Avimex and CONACYT.

## Introduction

Severe acute respiratory syndrome coronavirus 2 (SARS-CoV-2) emerged in China in late 2019 and has since then caused the coronavirus disease 2019 (COVID-19) pandemic (1, 2). Vaccines against SARS-CoV-2 were rapidly developed and have been shown to be safe and efficacious (3). However, in many low-and middle-income countries (LMICs) access to vaccines is still limited. In addition, mRNA based COVID-19 vaccines require frozen storage and transportation -severely restricting their usability in LMICs. Furthermore, production of many of the available COVID-19 vaccines is costly, affecting the price per dose. In addition, all currently approved COVID-19 vaccines are injected intramuscularly leading to strong systemic but absent or weak mucosal immunity (4) which is thought to be critical for achieving sterilizing immunity against SARS-CoV-2. Moreover, for more infectious variants like B.1.617.2 (Delta) the rate of breakthrough infections has increased (5) and has now peaked with the emergence of B.1.1.529 (Omicron) (6-13). These breakthrough infections are often asymptomatic or mild if symptomatic, and protection from severe disease remains high (14, 15). However, the fact that they occur is likely a consequence of the absence of persistent mucosal immunity, which can neutralize virus right at its entry point into the body, on mucosal surfaces of the upper respiratory tract. Vaccines that potentially induce mucosal immunity are better suited to induce sterilizing immunity and block transmission of a virus.

To address the issues raised above, we have developed a live Newcastle disease virus (NDV)-based SARS-CoV-2 vaccine. NDV is an avian paramyxovirus which is highly attenuated in mammals and has been tested in humans as an oncolytic virus and in preclinical models as a live vaccine vector (16-24). We engineered the LaSota vaccine strain of NDV to express the spike protein of SARS-CoV-2 (25-28). The version of the spike protein used is based on an enhanced immunogen design, which includes six proline mutations and a deletion of the polybasic cleavage site keeping the spike in a stable pre-fusion conformation (29). In addition, the ectodomain of the spike protein was grafted onto the transmembrane domain and cytoplasmic domain of the NDV fusion protein to ensure optimal incorporation into the Newcastle disease virion. The vaccine vector therefore carries spike on its surface and expresses it in cells that it infects.

NDV is an avian virus and it can be grown in embryonated chicken eggs to very high titers. Embryonated eggs are used for production of the majority of influenza virus vaccines used and therefore production capacity for this NDV-vectored vaccine already exists in high-income and LMICs (30). This also allows the vaccine to be produced at very low cost.

We have previously shown that an inactivated, as well a live version of this NDV-vectored vaccine, are safe, well tolerated, highly immunogenic and protective in animal models including in a swine model using different routes of administration, that contributed to the design of the phase I protocol reported herein (25-28, 31-33). Inactivated versions of the vaccine are currently in clinical development in Vietnam (NCT04830800), Brazil (NCT04993209) and Thailand (NCT04764422). Interim results from Thailand show that the inactivated formulation – which is injected intramuscularly -is safe and highly immunogenic (34). Here, we tested a live version of the vaccine, AVX/COVID-12-HEXAPRO (Patria, also known as NDV-HXP-S), in an open label non-randomized non-placebo-controlled phase I trial in 91 healthy volunteers. Vaccine was administered either via an intramuscular prime-boost regimen or, for optimal induction of mucosal immunity, via an intranasal prime-boost regimen. In addition, intranasal immunization followed by an intramuscular administration was also tested. Below, we report the interim safety and immunogenicity results from this trial in Mexico (NCT04871737).

## Methods

### Study design and participants

The phase I study was designed to evaluate the safety and immunogenicity of NDV-HXP-S given via three different vaccination strategies: intramuscular vaccination on day 0 and day 21, intranasal vaccination on day 0 followed by intramuscular vaccination on day 21 and intranasal vaccination on day 0 and day 21. In addition, three different dose levels were tested, 10^7.0^-10^7.49^ 50% egg infectious doses (EID_50,_ low dose (LD)), 10^7.5^-10^7.99^ EID_50_ (medium dose, MD) and 10^8.0^-10^8.49^ EID_50_ (high dose, HD), resulting in 9 groups with 10 participants each (**Table 1**). Female and male participants between 18 and 55 years of age without prior immunity to SARS-CoV-2 were enrolled. The protocol was designed by ProcliniQ Investigación Clínica, S. A. de C. V. with input from the Instituto Mexicano del Seguro Social (IMSS) and Laboratorio Avi-Mex, S.

**Table 1.** Distribution of subjects in groups per dose and administration route/regimen (IM=intramuscular; IN=intranasal)

| Dose | Administration route<br>1st Dose/2nd Dose |  |  |
| --- | --- | --- | --- |
|  | IM/IM | IN/IN | IN/IM |
| $10^{7.0}$ EID<sub>50</sub>/dose (LD) | 10 | 10 | 10 |
| $10^{7.5}$ EID<sub>50</sub>/dose (MD) | 10 | 10 | 10 |
| $10^{8.0}$ EID<sub>50</sub>/dose (HD) | 10 | 10 | 10 |

A. de C. V. (Avimex®), the later as sponsor with the statistical help of iLS Clinical Research, S. C. The study was approved by the Federal Commission for the Protection against Sanitary Risks (COFEPRIS) in Mexico under number 213300410A0063/2021, after approval by the Ethics, Biosafety and Research Committees of the clinical research site Hospital Medica Sur (03-2021-CI/CEI/CB-156) in full compliance of the Mexican regulation and under the principles of the Declaration of Helsinki and Good Clinical Practice. The samples for the immunological assays were processed at the National Institute for Respiratory Diseases (INER) in Mexico City.

The primary outcomes were to evaluate the safety of the three concentrations (viral titers) and three administration routes across nine groups. Immunogenicity measurements including the induction of IgM and IgG, neutralizing antibodies, cellular responses and induction of mucosal immunity (mucosal IgA, neutralizing IgA) were secondary outcomes.

### Study groups

This Phase I study was designed as a non-randomized open label study without placebo control group. Ninety volunteers were assigned to one of nine treatment groups in the order of enrolment according to **Table 1**. The first intervention of each treatment group was made sequentially to 18 sentinel subjects according to **Table 2**. The first 18 subjects (S1 to S18) received incrementally a dose from the lowest to the highest viral titer with no more than one subject per day, per dosage and route of administration. The safety data of the sentinel subjects was then evaluated by an independent Safety Data Monitoring Committee (SDMC) before authorizing the administration of the first vaccine dose to the rest of the subjects, who were then sequentially enrolled according to **Table 3**. The SDMC also evaluated the safety data of the full cohort after the first dose before the administration of the second dose to the nineteenth subject enrolled (first outside the sentinel group) on day 21 after the first dose.

**Table 2.** Incremental dose administered per route/regimen for the first 18 subjects as a sentinel group for safety monitoring (IM=intramuscular; IN=intranasal; S=subject)

| 1<sup>st</sup> Dose | Low Dose (LD)<br>($10^{7.0}$ EID<sub>50</sub>/dose) | | | Medium Dose (MD)<br>($10^{7.5}$ EID<sub>50</sub>/dose) | | | High Dose (HD)<br>($10^{8.0}$ EID<sub>50</sub>/dose) | | |
| --- | --- | --- | --- | --- | --- | --- | --- | --- | --- |
|  | Day1 | Day 2 | Day3 | Day 4 | Day 5 | Day 6 | Day 7 | Day 8 | Day 9 |
| IM | S1 | S3 | S5 | S7 | S9 | S11 | S13 | S15 | S17 |
| IN | S2 | S4 | S6 | S8 | S10 | S12 | S14 | S16 | S18 |
| Evaluation by an Independent Safety Committee 7 days after last High Titer vaccination |  |  |  |  |  |  |  |  |  |

**Table 3.** Assignment of first dose to subjects enrolled after sentinel group (IM=intramuscular; IN=intranasal; S=subject)

| 1<sup>st</sup> Dose | Low Dose (LD)<br>($10^{7.0}$ EID<sub>50%</sub>/dose) | Medium Dose (MD)<br>($10^{7.5}$ EID<sub>50%</sub>/dose) | High Dose (HD)<br>($10^{8.0}$ EID<sub>50%</sub>/dose) |
| --- | --- | --- | --- |
| IM | S19, S21, S23, S28, S30,<br>S32, S37 | S43, S45, S47, S52, S54,<br>S56, S61 | S67, S69, S71, S76, S78,<br>S80, S85 |
| IN | S20, S22, S24, S25, S26,<br>S27, S29, S31, S33, S34,<br>S35, S36, S38, S39, S40,<br>S41, S42 | S44, S46, S48, S49, S50,<br>S51, S53, S55, S57, S58,<br>S59, S60, S62, S63, S64,<br>S65, S66 | S68, S70, S72, S73, S74,<br>S75, S77, S79, S81, S82,<br>S83, S84, S86, S87, S88,<br>S89, S90 |
| Evaluation by an Independent Safety Committee 7 days after last HD vaccination |  |  |  |

There was a deviation with one of the subjects who reported negative results in the PCR and IgG/IgM tests for SARS-CoV-2 at screening, and who was therefore enrolled in the clinical trial and received the first intramuscular vaccine dose (Day 0) in the low dose (LD) group. However, a subsequent test of anti-spike antibodies, post-administration of the vaccine, showed a low, yet positive antibodies level (148.8 AU/mL, Elecsys® Anti-SARS-CoV-2 S, Roche Diagnostics). The investigators reviewed the case and considered that it was in the best interest of the subject to remain in the study, since the safety of the subject was not at risk and vaccination for the age group to which the subject belongs was at the time of the study not available under the Mexican national vaccination program. This decision would also be consistent with an ethical obligation of properly monitoring the safety of the volunteer. The subject consented to continue participation in the study with the sponsor’s authorization. Safety data was included in the safety analysis, but immunogenicity data from this subject was not considered for immunogenicity assessment.

For those subjects who received the first dose intranasally (IN), the second dose was administered by alternating the administration route. The first subject was given the second dose via the IN route followed by the second subject who was dosed by the IM route. This alternation continued until the IN/IN and IN/IM groups were dosed at each dose level according to **Table 1**. All subjects who received the first dose via IM also received the second dose via IM in order to complete the corresponding IM/IM groups.

As an additional circumstance around the protocol, it is important to stress that the study was conducted almost concurrently with a COVID-19 wave in Mexico driven by the emergence of the B.1.617.2 (Delta) variant in Mexico City (34). This circumstance affected the clinical trial as some of the participants were infected either between the first and the second dose or after the administration of the second dose as reported in **Table 4**.

**Table 4.** Subjects infected by SARS-CoV-2 per group (IM=intramuscular; IN=intranasal)

| Dose | Route/regimen<br>Infected after 1st Dose |  |  | Route/regimen<br>Infected after 2nd Dose |  |  | Total per<br>dose level |
| --- | --- | --- | --- | --- | --- | --- | --- |
|  | IM/IM | IN/IN | IN/IM | IM/IM | IN/IN | IN/IM |  |
| $10^{7.0}$ EID<sub>50%</sub>/dose (LD) | 1 | 0 | 0 | 0 | 0 | 0 | 1 |
| $10^{7.5}$ EID<sub>50%</sub>/dose (MD) | 0 | 2 | 0 | 0 | 3 | 1 | 6 |
| $10^{8.0}$ EID<sub>50%</sub>/dose (HD) | 1 | 1 | 0 | 0 | 0 | 1 | 4 |
| Subtotal per route and<br>administered doses | 2 | 3 | 0 | 0 | 3 | 2 |  |

According to the above, the total N for safety assessment was 91 participants and for immunogenicity assessments, the N was variable per group since subjects who acquired an infection (see **Table 4**) were excluded from analysis.

### Procedures

As mentioned above, AVX/COVID-12-Hexapro (Patria) is a Newcastle disease virus (NDV)-based SARS-CoV-2 vaccine based on the LaSota vaccine strain of NDV (25-27). It was engineered to express a version of the spike protein of SARS-CoV-2 which includes six proline mutations and a deletion of the polybasic cleavage site, keeping the spike in a stable pre-fusion conformation (29). In addition, the ectodomain of the SARS-CoV-2 spike was grafted onto the transmembrane and cytoplasmic domains of the NDV fusion protein to ensure optimal incorporation into the Newcastle disease virion. The vaccine was obtained as reported previously (25-27, 31) and manufactured under Good Manufacturing Practices at the COFEPRIS approved facilities of Laboratorio Avi-Mex, S. A. de C. V. in Mexico City. The vaccine was formulated without adjuvants in three different viral titers per dose (LD, MD, HD) as described above. For the intramuscular (IM) administration, it was formulated in single dose vials with the corresponding viral titer contained in 0.5 mL for administration as a single injection into the deltoid muscle. In the case of the intranasal (IN) administration, it was formulated in single dose vials as a 0.2 mL solution containing the corresponding viral titer, for administration of 0.1 mL in each nostril. The vaccines formulated as described were stored under refrigeration (4°C).

The 0.5 mL intramuscular dose was administered through a regular syringe and needle, and for the 0.2 mL intranasal route a nasal sprayer device coupled to the syringe (MAD Nasal™ - Intranasal Mucosal Atomization Device) was used instead.

The study was conducted at Hospital Medica Sur in Mexico City. A written informed consent was obtained from each participant, as approved by COFEPRIS, to voluntarily participate in the study for 12 months including 11 visits to the site plus at least six telephone follow-up calls scheduled according to the date of the first visit.

A screening visit was conducted 3 days before vaccination where each participant underwent a full medical history and examination. A medical history was obtained, including recording of all vaccines and medications received within the last 30 days, and daily activities that posed a high risk for getting infected with SARS-CoV-2. The physical examination included measurement of vital signs (blood pressure, heart rate, respiratory rate, and temperature), oxygen saturation, weight and height. At the screening visit participants were also subject to urine and blood testing, hematology, blood cell count, kidney and liver function test, blood lipids, and testing for human immunodeficiency virus (HIV), hepatitis B and C virus and syphilis, pregnancy tests for woman of childbearing potential, electrocardiogram, and thorax CT scan. In addition, the participants were subject to COVID-19 testing (nucleic acid-GeneFinder™ COVID-19 Plus RealAmpKit, and antibody-as above) to exclude prior or active infection, as such infection was part of the exclusion criteria. Further details on eligibility are provided in the trial protocol (**Appendix 1**).

Eligible subjects were enrolled and were administered the first vaccine dose corresponding to their group and given a patient diary at basal visit (D0). Vital signs were measured prior to the administration of each dose and at 90 minutes thereafter. Subjects were observed on-site during that period. Further daily telephone interviews were conducted from days 1 to 6 for collection of safety data and participants returned to the site on day 7 (D7) after the first dose administration (D0) followed by scheduled visits on days 14, 21, 28, 42, 90, 180 and 365. All on site visits included measurement of vital signs, weight, and determination of body mass index (BMI). Data for visits on days 90, 180 and 365 are not yet available since the trial is still ongoing.

Day 14, 21, 28, 42, 90, 180 and 365 visits include blood sampling for IgM – IgG – IgA antibodies, neutralizing antibodies, and T cell responses. In addition, those subjects who received at least one IN dose provided also saliva and nasal swab samples on these same dates. According to the study protocol, basal samples of saliva and nasal fluids were not collected as there was no previous infection and specific antibodies were likely negative.

A PCR test was also performed prior to the application of the second dose of AVX/COVID-12-Hexapro. As described above, participants positive for SARS-CoV-2 infection were considered for the safety assessment but excluded from the immunogenicity analysis.

Adverse events were documented based on standardized terms (MedDRA) and classified as Adverse Events (AE), Serious Adverse Events (SAE), both as defined by ICH/E6R2 Good Clinical Practices definitions. Adverse Events of Special Interest (AESI) were defined in the Protocol as those that appeared within 7 days from vaccination and categorized as local, when related to the injection or the intranasal administration (inflammation, redness, local increased temperature, itching, low-grade fever), or systemic or related to COVID-19 disease (fever, chills, cough, difficult breathing, muscular or articular pain, headache, anosmia, ageusia, odynophagia, nasal congestion or secretion, nausea or vomiting, diarrhea or fatigue).

The number and percentage of AEs, SAEs and AESIs were recorded after every vaccine administration. AESIs were considered associated with vaccination and evaluated 7 days after each vaccination, while AEs were assessed after 21 days as of vaccination. AEs intensity was generally registered as low, mild or severe according to the protocol. Clinically relevant abnormalities in laboratory tests or at physical examination were recorded by groups and then correlated to the vaccine viral titer and administration route.

## Immunological assay

### Sample collection

Blood samples, nasal exudates, and saliva samples were obtained as described above, according to the group, at the clinical research site and transported to INER at room temperature. Blood samples were processed within two and half hours of vein puncture. Biological samples were obtained before and 14, 21, 28 and 42 days after the first vaccination.

Venous blood was obtained using standard procedures and was collected into separator tubes (SST BD vacutainer tubes, Franklin Lakes, NJ, USA). Vacutainer tubes were centrifuged at 1200 rpm (centrifuge: Rotanta 460R; Rotor: 5624, Hettich, Tuttlingen DE) for 10 minutes to separate serum. The serum was then removed from the upper portion of the tube, aliquoted, and stored at -20°C until use.

Serum samples from convalescent individuals (N=51, collected at a median of 41 days post onset (standard deviation 12 days, range 21-65 days)) with SARS-CoV-2 infection confirmed by real-time reverse transcriptase PCR (RT-PCR) were collected after recovery on the day they resumed regular activities to be used as positive controls, and serum samples from healthy individuals obtained between 2014-2018 (prepandemic) were used as negative controls (INER approved protocol number B20-21and B22-12).

All blood samples and blood products, nasal exudates and saliva were handled in a BSL-2 laboratory with the use of appropriate personal protective equipment and safety precautions using processing protocols approved by the INER Institutional Biosafety Committee.

Venous blood was collected in sodium heparin tubes (BD vacutainer tubes, Franklin Lakes, NJ, USA) and diluted 1:1 within two and half hours for whole blood stimulation with 0.99 μg/mL of S1 subunit of the spike protein (RayBiotech, Peachtree Corners GA) in the presence of anti-CD28/CD49d (BD, San Jose CA) for 18h 20min at 37°C in 5% CO_2_.

### SARS-CoV-2-spike protein specific antibodies enzyme-linked immunosorbent assay (ELISA)

S1-specific IgG in serum samples were measured using two commercial kits from EuroImmun, following manufacturer’s instructions and using an analyzer (Euroimmun AG, Lübeck, Germany). Serum samples were diluted 1:100, and 100 μL of samples, calibrator, negative and positive control were added to each well and incubated at 37°C for 60 minutes. This step was followed by three washes using 300 μL of washing buffer per well. Then, 100 μL of the anti-human IgG, labeled with peroxidase were added and incubated at 37°C for 30 minutes for IgG detection. The plates were subsequently washed before the addition of 100 μL of substrate solution. After incubation for 30 minutes at room temperature, 100 μL of stop solution was added and the optical density (OD) was read at 450 nm in the analyzer (EuroImmun) within 30 minutes after adding the stop solution. The results were reported as the ratio between the extinction of the sample and the extinction of the calibrator, and a ratio of > 1.1 was considered positive (35). For serum analysis, twofold serial dilutions of sera were processed as described above, and the end-point titer was calculated as the most diluted serum concentration that gave a ratio >1.1. The limit of detection was 1:100, samples with activity below the limit of detection were assigned a titer of 1:50 for graphing purposes. Samples ran across multiple plates were calibrated using a manufacturer-provided calibrator solution.

### Receptor binding domain (RBD) – angiotensin converting enzyme 2 (ACE2) interaction inhibition assay (RAIIA)

To determine the presence of antibodies that block interaction between the spike receptor binding domain (RBD) and the angiotensin converting enzyme 2 (ACE2) receptor, we used a commercial assay from GenScript, which is a protein-based surrogate neutralization assay (36). Samples were analyzed following the manufacturer’s instructions (GenScript version RUO 3.0 update 01/02/2021). Briefly, samples and controls were diluted 1:10 in kit sample buffer and mixed 1:1 with horseradish peroxidase (HRP)-conjugated recombinant SARS-CoV-2 RBD fragment (HRP-RBD) and incubated at 37°C for 30 minutes to allow binding of circulating antibodies to HRP-RBD. The mixture was then added to the capture plate which is pre-coated with the human ACE2 protein. Unbound HRP-RBD as well as any HRP-RBD bound to non-neutralizing antibody was captured on the plate, while circulating neutralization antibodies-HRP-RBD complexes remained in the supernatant and get removed during washing. Then 3,3’,5,5’-tetramethylbenzidine (TMB) solution was added. By adding stop solution, the reaction was quenched and the plates were read at 450 nm using Analyzer 1 (EuroImmun). Absorbance of a sample is inversely correlated with blocking RBD-ACE2 interactions. The results are expressed as the percentage (%) of inhibition and 30% inhibition was used as cutoff as previously established (36).

### Intracellular cytokine staining assay

Whole blood diluted 1:1 was stimulated with RPMI 1640 medium (Lonza, Walkersville, MD) 0.99 μg/mL of S1 subunit of spike protein in the presence of anti-CD28/CD49d (BD, San Jose CA) for 18 h20 min at 37°C in 5% CO_2_. GolgiStop (BD, San José, CA) was added, and the samples were cultured additionally for 4 h. Medium was used as a negative control and 10 μg/mL of PHA (Sigma-Aldrich) as a positive control. Samples were washed with phosphate buffered saline (PBS) without Ca^2+^ and Mg^2+^ (Lonza), and stained with Live/Dead near-IR Dead Cell Stain Kit for 633 or 635nm excitation (Invitrogen, Eugene, OR), for 15 min at room temperature in the dark. Then red blood cells (RBCs) were lysed with RBC lysis buffer (BD) for 10 min followed by a washing step with staining buffer PBS without Ca^2^+ and Mg^2^+ supplemented with 1% fetal bovine serum (FBS) and 0.1% NaN_3_). Cell surface staining was performed using a cocktail of anti-human CD3, CD4 and CD8 antibodies in staining buffer for 15 min at room temperature in the dark. After an additional washing step with staining buffer, the cells were fixed and further permeabilized using BD Cytofix/Cytoperm following the manufacturer’s instructions. Intracellular staining was performed in Cytoperm using a anti-human IFN-γ antibodies for 30 min at room temperature in the dark. Cells were washed with BD Perm/Wash buffer and further resuspended in PBS. Cells were kept at 4°C in the dark until acquisition and analysis. Unstained and fluorescence minus one (FMO) controls were included. Details of the antibodies used in the flow cytometry assay are listed in Supplementary Table 1, and the rest of the reagents in Supplementary Table 2. At least 200,000 events of the lymphocyte region in a forward scatter (FSC) vs side scatter (SSC) scatter plot were acquired in a fluorescence-activated cell sorter (FACS) Aria II (BD). Analysis was performed using FACS Diva 8.0. The gates applied for the identification of SARS-CoV-2 antigen-specific cytokine-producing CD3+, CD4+ or CD8+ cells were defined using the FMO controls and used for limit of detection (LOD) (37).

## Outcome

### Primary outcomes

Primary outcomes of the study were established as follows:

- To evaluate the safety of three concentrations (10^7.0-7.49^, 10^7.5-7.99^, 10^8.0-8.49^ EID_50%_/dose) of the recombinant vaccine against SARS-CoV-2 based on a Newcastle Disease virus (rNDV) administered twice intramuscularly, twice intranasally or intranasally followed by intramuscularly in healthy volunteers

### Secondary outcomes

- To evaluate the immunogenicity of three concentrations (10^7.0-7.49^, 10^7.5-7.99^, 10^8.0-8.49^ EID_50%_/dose) of the recombinant vaccine against SARS-CoV-2 based on a Newcastle Disease virus (rNDV) administered twice intramuscularly, twice intranasally or intranasally followed by intramuscularly in healthy volunteers
- To evaluate the nasal mucosal humoral immunity of three concentrations (10^7.0-7.49^, 10^7.5-7.99^, 10^8.0-8.49^ EID_50%_/dose) of the recombinant vaccine against SARS-CoV-2 based on a Newcastle Disease virus (rNDV)

This manuscript describes an interim analysis which focuses on initial safety data and binding and ACE2/RBD interaction inhibiting antibodies and T-cell based immunity. Other readouts will be described in future publications.

### Statistical analysis

Interim analyses were scheduled for days 21, 28, 42, and after 6 and 12 months (end of study). This report includes data obtained up to day 42. For continuous variables, one-way ANOVA and Student t test were used, and non-parametric tests were used for discrete (count) variables. Safety endpoints were expressed as frequencies (%) with 95% binomial exact confidence intervals (Cis), while immunological assessment are expressed as median and IQRs. In this report all analyses are descriptive only, as samples are still pending further analyses and the results reported here are preliminary in nature. IgG titers are reported per group as geometric mean titers (GMT) with a 95% CI at days 0 (basal), 14, 21, 28 and 42. For logarithmically transformed antibody titers ANOVA and Wilcoxon’s rank sum test were used for data not distributed normally, and significances between groups were paired and differences assessed with a 95% CI. Bilateral CIs for GMT were calculated by back-transformed 95% CI based on Student t tests for titers with log_10_ transformation. The proportion of subjects with a titer above a predetermined parameter for IgG, IgM and IgA with 95% CI at days 14, 21, 28, and 42. Days 90, 180, as well as 6 and 12 months will also be analyzed after completion of the study. Seroconversion rate and 95% CI with respect to the basal titer was also determined as the proportion of subjects with detected titers of specific antibodies for the spike protein of SARS-CoV-2 as determined by ELISA and an ANOVA with 95% CI was used for assessing the capacity of circulating antibodies to inhibit the interaction between RBD and ACE2. T cell mediated responses were assessed as a proportion of positive responders through a χ2 test or Fisher’s exact test for categorical data. Finally, the titers of antibodies per administration form (IM/IM, IN/IN and IN/IM) were compared using two-way ANOVA. The full statistical analyses details are provided in the trial protocol (**Appendix 1**), and will be performed in full upon completion of all procedures.

### Role of funding source

The funding for the clinical study was provided by the National Council for Science and Technology (CONACYT, México), except for all the production and vaccine product supply which was funded solely by Avimex. CONACYT did not participate in the trial design but did evaluate it and approved the project through their National Committee on Research, Development and Innovation on Public Health. Funding was managed by Avimex and used to pay for all laboratory tests, clinical site and clinical professionals. CONACYT also facilitated the identification, purchase and importation of certain supplies and the communication with other entities of the Federal Mexican Government to facilitate the study.

## Results

From May 24^th^ 2021 to August 20^th^ 2021, 153 volunteers were assessed. Two voluntarily withdrew from the study, while 49 were excluded as they did not meet eligibility criteria. 91 volunteers were enrolled into the nine different groups and either dosed twice IM (IM-IM groups), dosed sequentially IN followed by IM (IN-IM groups) or received two IN vaccinations (IN-IN). groups) in a three week interval (**Figures 1 and 2, Tables 1-3**). Three different dose levels, low dose (LD), medium dose (MD) and high dose (HD) were evaluated. Distribution of participants by gender between the safety population groups did not show statistically significant differences according to the dose/route of administration. All the participants identified themselves as Mestizo. Regarding distribution of patients according to age, there were no significant differences either between groups that received low, medium, or high doses by any administration routes. Average ages, age range of participants, gender distribution, weight, height, and body mass index in each study group are indicated in **Figure 1C**. Up to day 45 post first vaccination none of the enrolled individuals were excluded from the study for safety evaluation but one subject had to be excluded from the immunogenicity evaluation due to a positive baseline titer and several subjects had to be excluded due to SARS-CoV-2 infections (**Table 4**).

**Figure 1.**
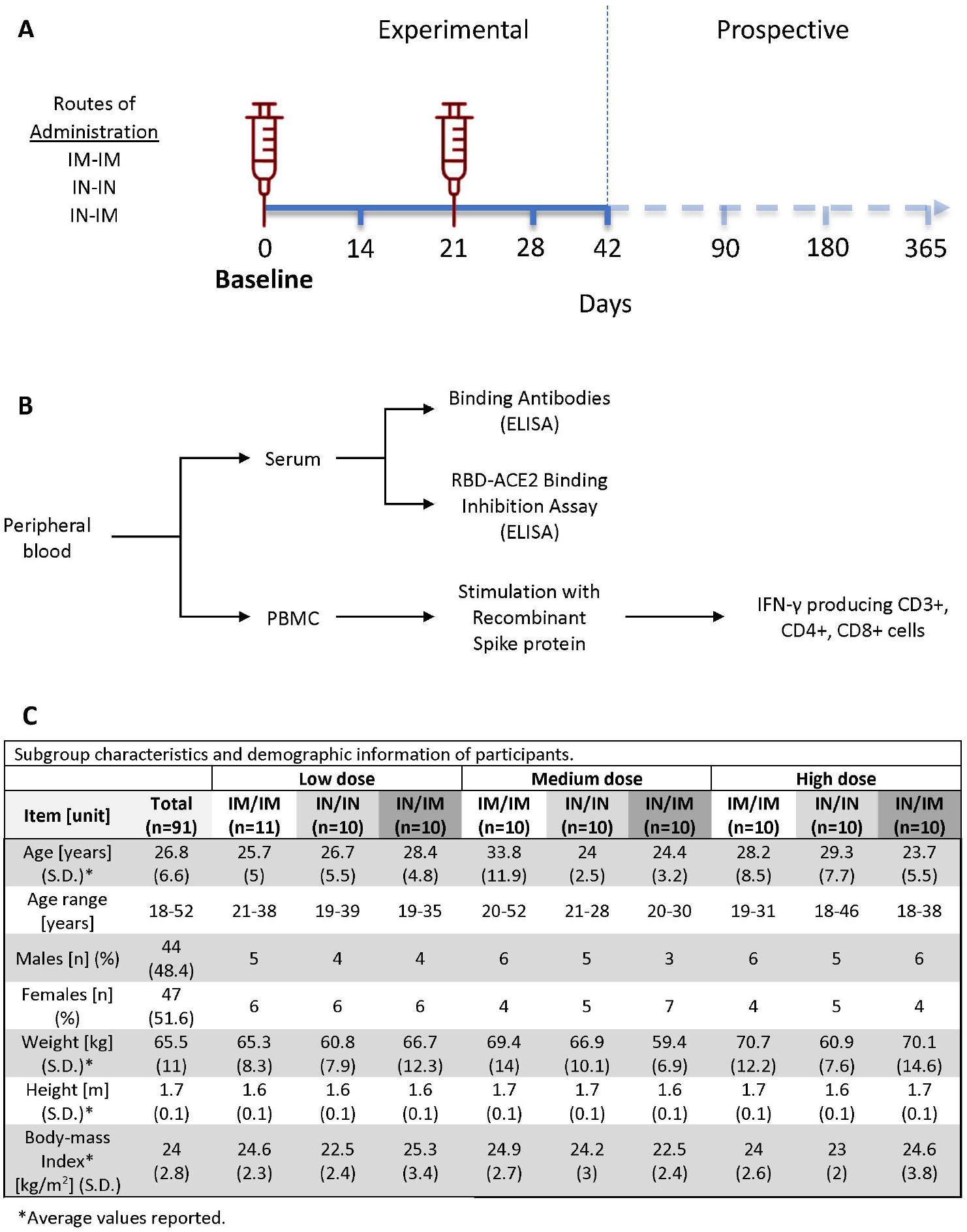
Study design and groups distribution. (A) Schematic representation of the study timeline, indicating routes of administration, vaccination time points, and sample collection for immunogenicity analyses. The three different vaccination regiments tested; intramuscular (IM) followed by intramuscular (IM), intranasal (IN) followed by intranasal (IN), and intranasal (IN) followed by intranasal (IN) administration are show on the left. Time points of sample collection (0, 14, 21, 28, 42, 90, 180 and 365 days after the first vaccine dose administration) and time points of vaccine administration (indicated by the red syringe) are shown on the right. (B) Diagram depicting specimen types collected to assess immunogenicity. (C) Subgroup characteristics and demographic information of participants of the trial (n=91).

**Figure 2.**
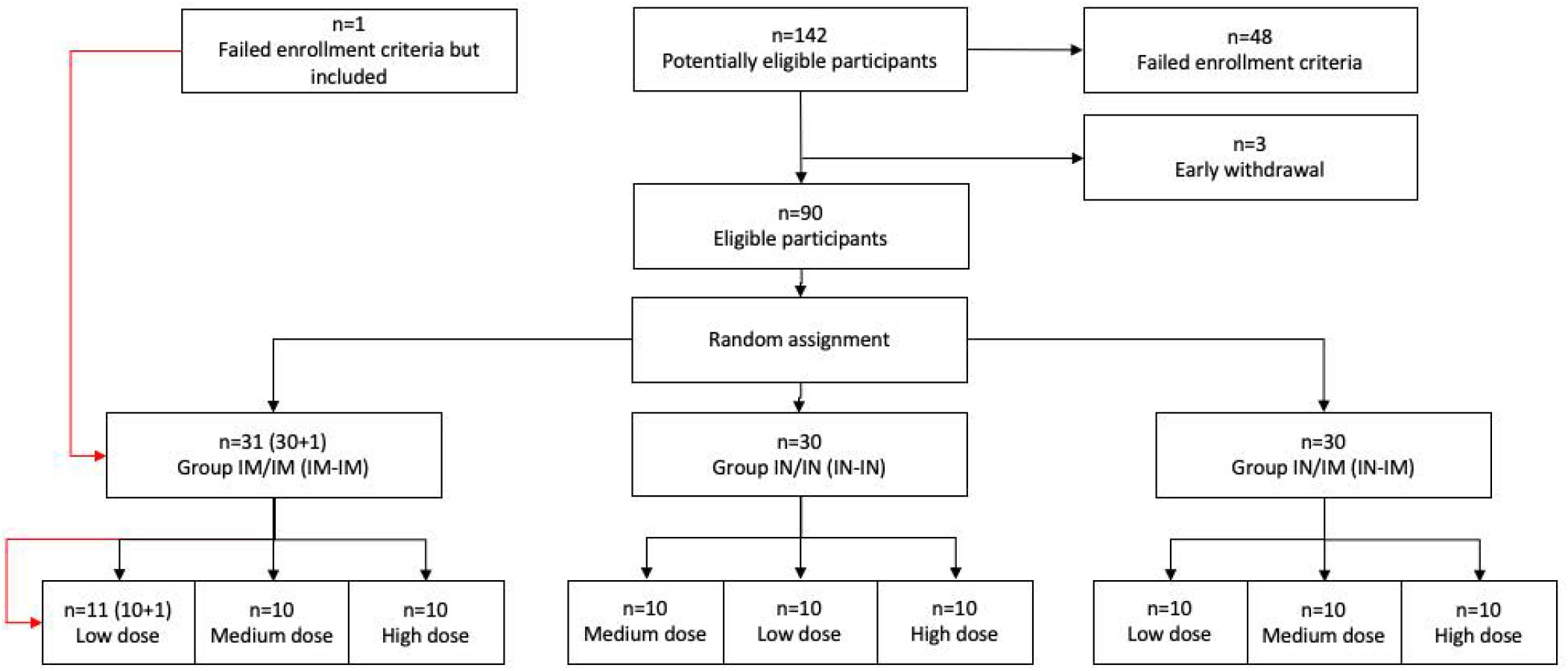
Enrollment and sub-randomization. Diagram representing number of participants initially screened (n=142), failed enrollment criteria (n=48), early withdrawals (n=3), and eligible participants (n=90) that were included in the trial and assigned to any of the three vaccination regimens (n=30, per group) and dose (low n=10, medium n=10, high=10). A participant that initially was considered eligible and received an IM-IM regimen, but subsequently failed study criteria is indicated on the left.

In general, all formulations were well tolerated with little reactogenicity detected (**Figure 3**). Up to day 45 post first vaccination of the latest enrollment of a subject, there had been 625 adverse events in total, of which 319 occurred within the period considered as of special interest (within 7 days after either of the two administrations). Of these 319 events within the special interest period, 66 were considered local and 253 systemic. In general, the distribution of AEs among the different groups of the study did not present statistically significant differences in terms of the number of individuals who presented at least one event, nor differences in the incidence of events according to their severity, except for those of the IN route which of course did not show the injection-related AEs. Additionally, none of the routes of administration or dose evaluated were associated with serious adverse events.

**Figure 3.**
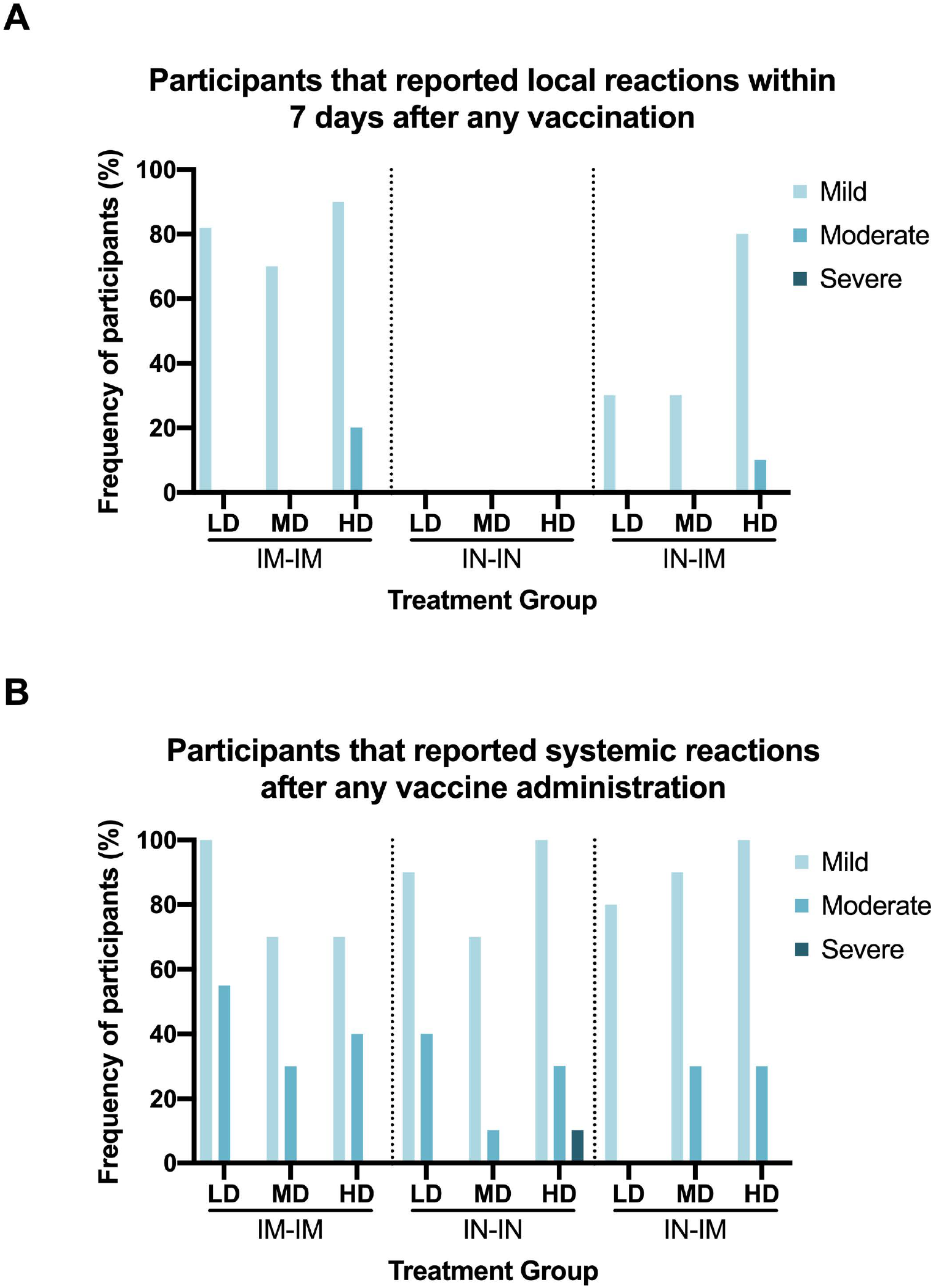
Local and systemic solicited adverse reactions. Adverse events (AE) were registered according to the standardized MedDRA dictionary terms and classified as mild, moderate, or severe according to ICH/E6R2 Good Clinical Practices definitions. (A) Adverse reactions reported by the trial participants within 7 days after the first and second vaccine doses are shown. (B) Systemic reactions reported by the trial participants throughout the observational period are shown. In both A and B, mild, moderate, and severe adverse reactions are shown in the individuals receiving either of the three vaccination regimens and data is stratified by the vaccine dose received (LD=low dose, MD=medium dose, and HD=high dose).

Out of 625 adverse events, 552 (88.32%) were of mild intensity, 68 (10.88%) moderate and only 5 (0.8%) severe intensity events were recorded. Distribution of adverse events between the different groups by route of administration or by dose received was not statistically significant. No deaths or significant/serious adverse events were reported, and no alterations of vital signs or clinically significant events were reported.

To determine immunogenicity of the different vaccine doses and vaccination routes we first performed ELISAs against the S1 domain of the spike protein (**Figure 4**). S1 was chosen as target because this subdomain of the spike includes the N-terminal domain and the RBD, which host most of the neutralizing epitopes. In addition, a reliable commercial ELISA focusing on that target was available locally. For the IM-IM vaccination regimen, little induction of anti-S1 antibody was observed after the first dose. However, the second dose boosted titers in a dose dependent manner with high reactivity in the HD group and lower reactivity in the MD and LD groups. As expected, the response after IN vaccination was lower and substantial reactivity was only detected in the HD group post-boost with 56% of the individuals in the group having detectable titers. Finally, in the IN-IM regimen, the reactivity was similar to the IM-IM regimen with an 89% response rate after the boost. Antibody titers induced by the HD IM-IM and IN-IM regimens were in general comparable or higher than the titer of convalescent individuals.

**Figure 4.** Spike-reactive antibody levels in sera from vaccinated volunteers. Antibodies against the S1 subunit of the spike protein (which contains the receptor binding domain (RBD)) were measured in vaccinees’ sera by ELISA at baseline, and 14, 21, 28, and 42 days after the first vaccine dose administration. Individuals receiving the IM-IM regimen (left column), IN-IN regimen (middle column), or IN-IM (right column), with a high dose (top row), medium dose (middle row), or low dose (bottom row) of the vaccine are shown. Human convalescent serum (HCS) samples were added as additional controls. The limit of detection (LoD) is indicated by the horizontal dotted line. Negative values are indicated as half of the LoD. Statistical significance is indicated as follows: *P < 0.05, **P < 0.01, ***P < 0.001.

While binding antibodies are important indicators of immunogenicity and have been correlated with protection (38, 39), we also wanted to determine functional antibody titers. A neutralization assay was unavailable for this interim analysis but we performed a surrogate assay, which measures inhibition of the interaction between the RBD and ACE2 (36). The titers detected in this assay do reflect results from the binding assay (**Figure 5**). For the HD IM-IM group the first vaccination increased the inhibitory titer just slightly. However, strong inhibitory activity was observed at post boost time points. This was also observed in the MD and LD groups, although more variability was detected there. For the IN-IN groups little inhibitory activity was detected and only in the HD group subjects. The IN-IM groups showed an intermediate phenotype with all individuals in the HD group having post-boost inhibitory antibodies. The response rate in the MD group was lower and only 20% of individuals in the LD group had activity above the limit of detection. Inhibition in the HD IM-IM regimen was in general comparable to inhibition of convalescent individuals. Of note, this assay does not allow to determine differences between groups with very strong responses. To summarize the binding and inhibition data, an analysis of the frequency of individuals with detectable binding or inhibiting antibody titers was performed (**Supplementary Figure 1**). High frequencies of subjects with binding and inhibiting antibodies were present in the all-dose groups receiving an IM-IM regimen, while moderate to high frequencies were detected in the HD IN-IM group.

**Figure 5.**
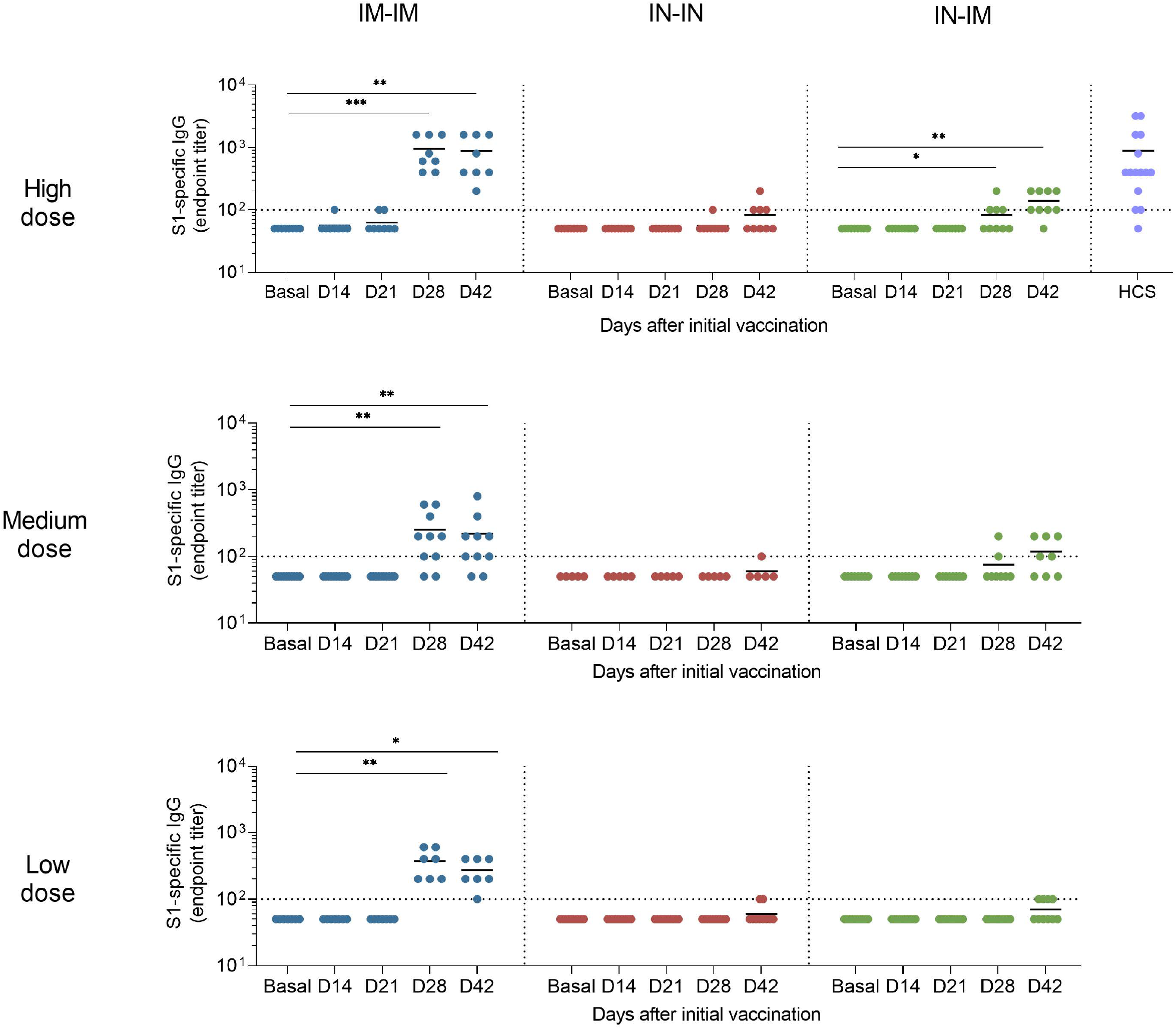
RBD-ACE2 interaction inhibiting antibodies in sera from vaccinated volunteers. Antibodies binding to the receptor binding domain (RBD) that inhibited its interaction with the angiotensin-converting enzyme 2 (ACE2) were assessed in vaccinees’ sera using an RBD-ACE2 interaction inhibition assay at baseline and 14, 21, 28, and 42 days after the first vaccine dose administration. Individuals receiving the IM-IM regimen (left column), IN-IN regimen (middle column), or IN-IM (right column), with a high dose (top row), medium dose (middle row), or low dose (bottom row) of the vaccine are shown. Human convalescent serum (HCS) samples were added as additional controls. The cutoff established for positivity (30%) in this assay is indicated by the horizontal dotted line. Statistical significance is indicated as follows: *P < 0.05, **P < 0.01, ***P < 0.001, ***P < 0.0001.

Cellular immune responses have been shown to be important for protection from SARS-CoV-2 infection, especially when neutralizing antibody titers are low (40). Here we assessed specific cellular immune responses by determining the percentage of CD3+, CD4+ and CD8+ cells that produced interferon γ (IFN γ) upon stimulation with the spike protein. A significant induction of IFN producing CD3+ cells was detected in all three HD vaccination regimens but not in the MD and LD groups when comparing day 42 with day 0 (**Figure 6**). While a trend was seen for IFN producing CD4+ cells in the HD IM-IM and IN-IN groups the increase was only statistically significant for the IN-IM HD group. No significant increases were found for CD4+ in the MD and LD groups. For CD8+ IFN producing cells a trend was also observed for the IM-IM HD group and the induction was significant for the HD IN-IN and IN-IM groups but not for any of the MD and LD groups. As a control of the specificity of the assay, a comparison of medium vs spike stimulated cells was performed (**Supplementary Figure 2**). Most of the participants had undetectable levels of activated CD3+ T cells upon stimulation with medium.

**Figure 6.**
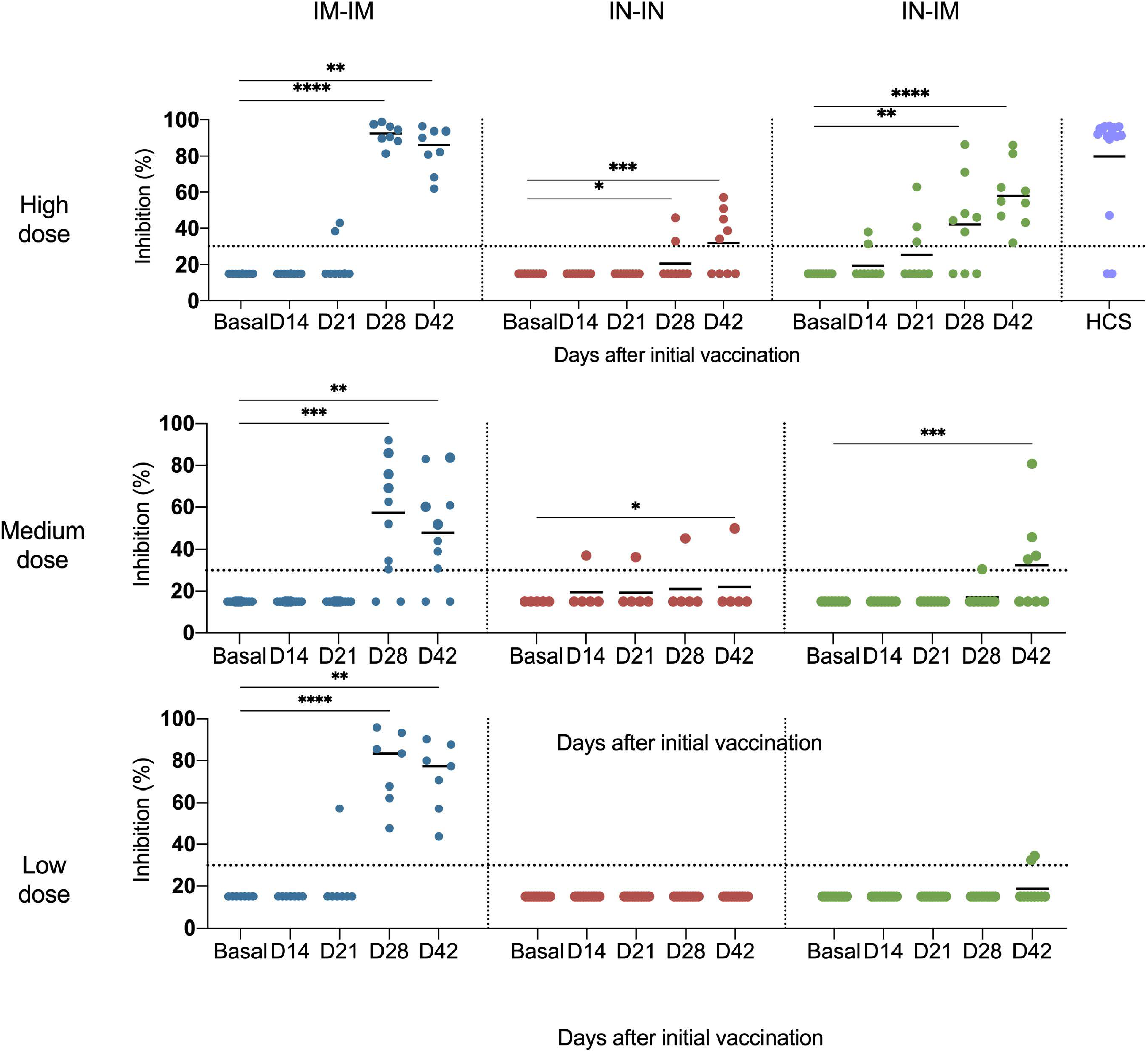
Activation profile of spike-specific CD3+, CD4+ and CD8+ T cells after vaccination. PBMCs were collected from vaccinees at baseline and 42 days after the first vaccine dose administration. Individuals receiving the IM-IM regimen (left column), IN-IN regimen (middle column), or IN-IM (right column) stratified by vaccine dose received (low, medium, or high) are shown. Activated CD3+ (top row), CD4+ (middle row), and CD8+ (bottom row) T cells were determined by flow cytometry after 18 h incubation with recombinant spike protein. Frequencies of T cells producing interferon gamma (IFN-γ) are presented. Statistical significance is indicated as follows: *P < 0.05, **P < 0.01.

**Figure 7.**
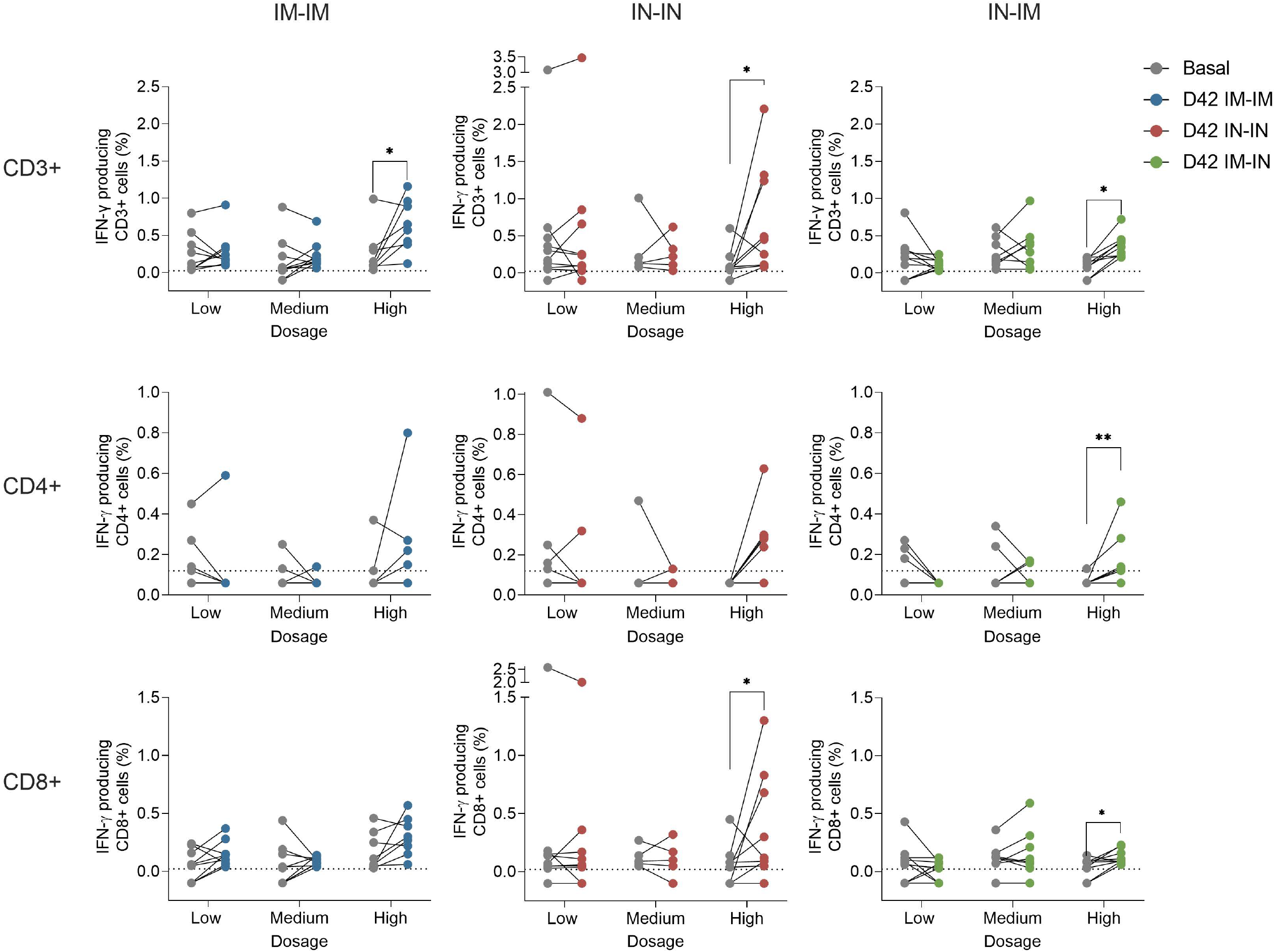

As described above, breakthrough infections did happen during the clinical trial. At day 42, there were 10 cases detected among groups, with no apparent trend dependent on dose or administration route (**Table 4**). The 10 cases were symptomatic, symptoms were mild, and none required hospitalization. 50% of the cases occurred before the second dose and the other 50% of the cases occurred after the second dose.

Assessment of safety and immunogenicity will continue for 12 months with sampling for immunogenicity planed at the 90-, 180-, and 365-day time points.

## Discussion

NDV-HXP-S can be produced at low cost and large scale using traditional egg-based influenza virus production processes. Influenza virus production capacity is available globally in high-income countries but also in LMICs (30). In addition, veterinary vaccine producers often also have egg-based production capacity which can be adapted for good manufacturing practice (GMP) production of human vaccines. The development of AVX/COVID-12-HEXAPRO could therefore alleviate the unmet global need for additional COVID-19 vaccine doses. Importantly, the superior spike antigen design of the NDV-vectored SARS-CoV-2 vaccine (29) is an additional advantage. Also, the favorable reactogenicity profile (as described here and in (41)), which is akin to that of influenza virus vaccines, makes the NDV-based vaccine likely more tolerant than mRNA or adenovirus-vectored vaccines (42-45). Furthermore, as demonstrated here, live NDV-vectored SARS-CoV-2 vaccine can be administered via the IN route. While assessment of mucosal immunity is not part of this interim report, intranasal vaccination is known to induce mucosal immunity that can potentially lead to sterilizing immunity and complete block of transmission.

Here we demonstrated that administration of live of AVX/COVID-12-HEXAPRO is safe and well tolerated at all dose levels. However, only the HD vaccine regimen induced robust antibody and cellular immune responses when given via the IM-IM or IN-IM routes, comparable to those in convalescent individuals. Cellular immune responses were induced by the IN-IN route but systemic antibody responses were not as robust. These results mirror those obtained with live AVX/COVID-12-HEXAPRO in the pig and rat models (31, 33). Given the robust immunogenicity and the high tolerability of the HD IM-IM and IN-IM vaccination regimens, it is justified to further develop these two modalities in Phase II trials. Importantly, given the high seroprevalence for SARS-CoV-2 in many regions globally and given the need for booster doses in a part of the population (elderly, immunosuppressed, health care workers etc.), the HD vaccination regimens should certainly also be evaluated in individuals with pre-existing immunity, likely as single IM or IN administrations. Currently, a single-dose booster trial is ongoing in Mexico City based on the data reported here with one IM and IN HD, and a phase II/III Study based on the HD IM-IM scheme is about to start in Mexico.

Our study has several limitations. Quantitative neutralization assays with authentic SARS-CoV-2 could not be performed at the study site at the time of analysis due to biosafety restrictions. In addition, in this interim analysis, neutralizing activity against variants of concern could not be assessed. Furthermore, we were not able to directly compare immune responses induced by inactivated NDV-vectored SARS-CoV-2 vaccines (41) with those observed following administration of other authorized/licensed COVID-19 vaccines. We expect to perform these additional assays and direct comparisons at later time points as soon as reagents and materials become available. So far, the assessment of mucosal antibodies has also not been possible. Finally, this was a non-randomized open label study without a placebo control group, which is more prone to biases as compared to randomized and double-blinded study designs.

In conclusion, we show that the live AVX/COVID-12-HEXAPRO vaccine has a safety profile that is remarkably independent of the dose and administration route with low frequency and intensity. Furthermore, the HD IM-IM and IN-IM vaccination regimens showed strong evidence of immunogenicity warranting further development of this vaccine candidate. Finally, it is important to note that the NDV vector technology is amenable to rapid changes in antigens expressed allowing for strain changes to match emerging viral variants. A B.1.1.529 (Omicron)-specific version of NDV-HXP-S is currently in development.

## Data Availability

All data produced in the present study are available upon reasonable request to the authors.

## Acknowledgements

The Mexican Government supports Patria via CONACYT. Its General Director and its Deputy General Director on Technological Development, Cooperation and Innovation, respectively Drs. Maria Elena Alvarez-Buylla Roces and Delia Aideé Orozco Hernández, have been directly in charge of inter-institutional liaison, and overall facilitation of proceedings, committee evaluations, sanitary set up and supervision, as well as of trial design approvals, evaluations and progress.

S.P.-R, J.J.C-M, P. S-D, Y.L-V and A.M. contributed to this study pro-bono and declare no competing interests.

We additionally acknowledge the broader support from various teams within Universidad Nacional Autónoma de México (UNAM) and Instituto Mexicano del Seguro Social (IMSS).

From Laboratorio Avi-Mex, S. A. de C. V., we acknowledge the following people for their operative support: Bernardo Lozano Alcántara, Carlos Woolfolk Frias, Leticia Espinosa Gervasio, Rodrigo Yebra Reyes, Vanessa Escamilla Jiménez, Juan Pablo Robles Alvarez, Avirán Almazán Gutiérrez, and Guadalupe Aguilar Rafael.

From INER, we acknowledge the following people for their technical support: Liliana Figueroa Hernandez, Francisco Cruz Flores, Lizeth García Cisneros, Claudia Ivett Hernandez Lázaro, María Angélica Velázquez González, Damaris Romero Rodriguez, Jessica Romero Rodriguez, Dulce Cinthia Soriano Hernández, Horacio Zamudio Meza, and Milton Nieto Ponce.

From ProcliniQ we acknowledge the support from Enrique Camacho-Mezquita, Juan Francisco Galán-Herrera and Mariana López-Martínez.

The salary of PP was partially funded by NIH (Centers of Excellence for influenza Research and Response, 75N93021C00014), U.S. NIAID grant (P01 AI097092-07), U.S. NIAID grant (R01 AI145870-03), by the NIH Collaborative Influenza Vaccine Innovation Centers contract 75N93019C00051 and a grant from an anonymous philanthropist to Mount Sinai. Design and generation of reagents used in preparation for this project in the Krammer laboratory were supported by Centers of Excellence for influenza Research and Response (75N93021C00014) and Collaborative Influenza Vaccine Innovation Centers (75N93019C00051), as was the García-Sastre laboratory. The authors thank Dr. Randy Albrecht for management of import/export of Newcastle disease virus vectors at the Icahn School of Medicine at Mount Sinai.

## Conflict of interest statement

PP reports financial support from the U.S. NIAID (Centers of Excellence for Influenza Research and Response 75N93021C00014, P01 AI097092-07, R01 AI145870-03). AGS reports financial support from the U.S. NIAID (Centers of Excellence for Influenza Research and Response 75N93021C00014, Collaborative Influenza Vaccine Innovation Centers contract 75N93019C00051). FK reports financial support from the U.S. NIAID (Collaborative Influenza Vaccine Innovation Centers contract 75N93019C00051, Center of Excellence for Influenza Research and Surveillance contract HHSN272201400008C), the JPB Foundation and the Open Philanthropy Project (research grant 2020-215611, 5384), and the U.S. NCI (contract 75N91019D00024, task order 75N91020F00003); he also has received royalties (Avimex), consulting fees (Pfizer, Seqirus, Third Rock Ventures and Avimex), and payment for academic lectures during the past two years. The NDV construction administered in this study was developed by faculty members at the Icahn School of Medicine at Mount including WS, PP, AGS, and FK. Mount Sinai has filed patent applications related to SARS-CoV-2 serological assays and the NDV-based SARS-CoV-2 vaccines; the institution and its faculty inventors could benefit financially. The live vaccine used in the study was developed by members of Avimex and Avimex filed patent applications with Mount Sinai and CONACYT. M.T-R., D.S.-M., E.S.P., C.R.L-M., H.E.C-C, F.C-P., G.P-DLR and B.L-D. are named as inventors on at least one of those patent applications. The clinical study was entirely performed in Mexico. The rest of the participants are employees of their corresponding institutions.

## Tables

**Supplementary Table 1.** Flow Cytometry antibodies.

| Antibody | Fluorochrome | Clone | Dilution | Brand | Catalog<br>Number |
| --- | --- | --- | --- | --- | --- |
| Anti-human IFN-γ | FITC | 4S.B3 | 1:200 | BD | 554551 |
| Anti-human CD3 | Alexa Fluor<sup>®</sup> 700 | SK7 | 1:33 | BioLegend | 344822 |
| Anti-human CD4 | PerCP/Cy5.5 | RPA-T4 | 1:100 | BioLegend | 300530 |
| Anti-human CD8 | PE/Cy7 | 53-6.7 | 1:100 | BioLegend | 980910 |

**Supplementary Table 2.** Flow Cytometry reagents.

| Reagent | Brand | Catalog Number |
| --- | --- | --- |
| Live/Dead Fixable near 633 or 635nm<br>(work dilution 1:1000) | Invitrogen | L34976A |
| Anti-CD28/CD49d | BD | 347690 |
| Phytohemagglutinin (PHA) | Sigma |  |
| Recombinant Spike Protein, Subunit 1 | Raybiotech | 230-011101-500 |
| RBC Lysis Buffer | Biolegend | 420302 |
| Golgi Stop | BD | 554715 |
| Cytofix/Cytoperm | BD | 51-2090KZ |
| Perm/Wash | BD | 51-2091KZ |
| PBS | Lonza | 17-516Q |
| RPMI 1640 | Lonza | 12-16Q |

## Figure legends

**Supplementary figure 1.**
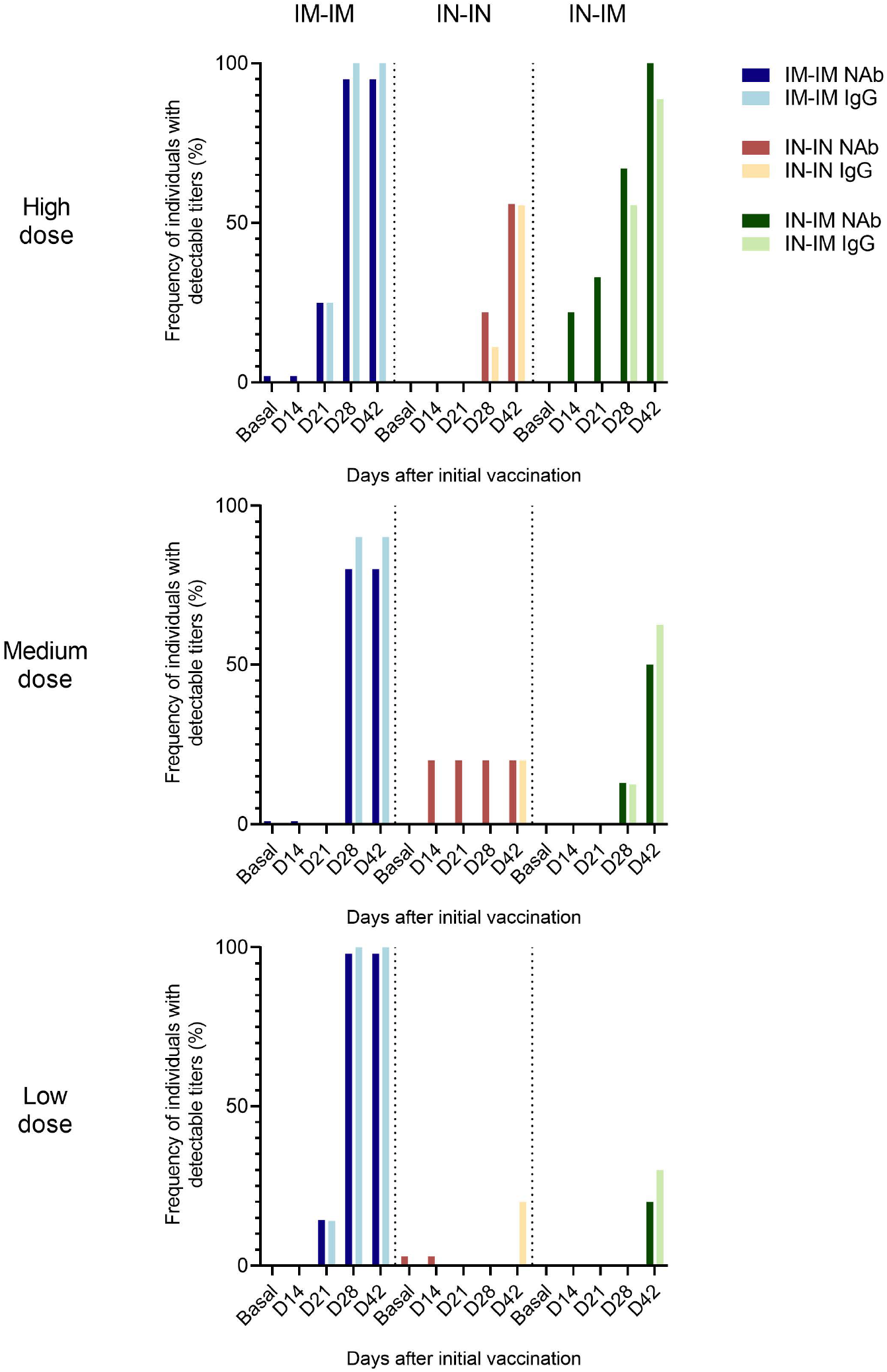
Frequency of individuals with detectable spike-reactive and RBD-inhibiting antibody titers. Antibodies against the S1 subunit of the spike protein (which contains the receptor binding domain (RBD)) and antibodies binding to the RBD that inhibited its interaction with the Angiotensin-Converting Enzyme 2 (ACE2) were assessed in vaccinees’ sera at baseline and 14, 21, 28, and 42 days after the first vaccine dose administration. Individuals receiving the IM-IM regimen (left column), IN-IN regimen (middle column), or IN-IM (right column), with a high dose (top), medium dose (middle), or low dose (bottom) of the vaccine are shown. S1-IgG = antibodies binding to the S1 spike subunit; NAb = antibodies inhibiting RBD-ACE2 interactions.

**Supplementary figure 2.**
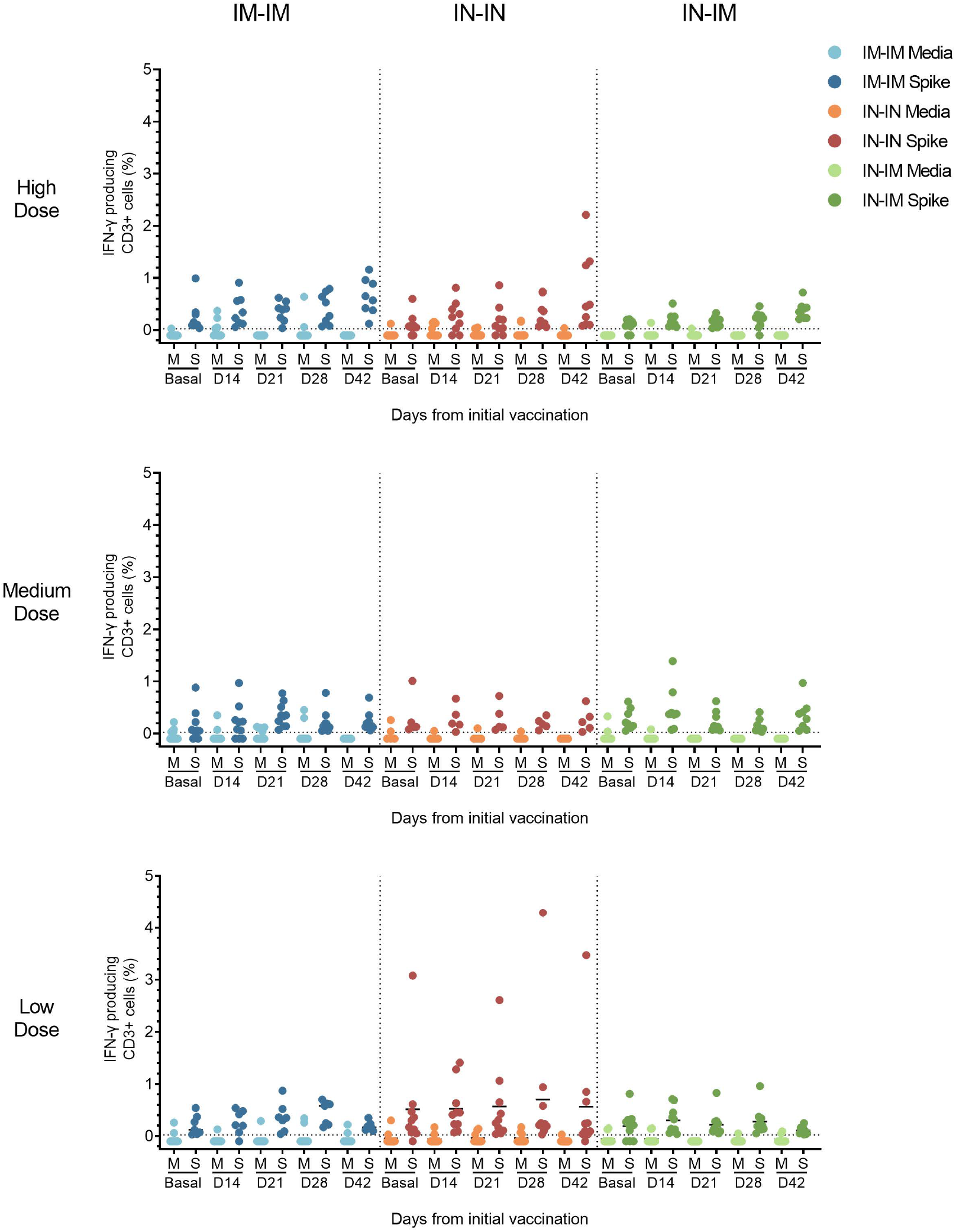
Medium vs antigen stimulation of CD3+ T cells from vaccinated volunteers. PBMCs were collected from vaccinees at baseline, and 14, 21, 28 and 42 days after the first vaccine dose administration. Individuals receiving the IM-IM regimen (left column), IN-IN regimen (middle column), or IN-IM (right column) stratified by the vaccine dose received (high, medium, or low) are shown. Activated CD3+ T cells were determined by flow cytometry after 18 h incubation with the recombinant spike protein or with medium only. Frequencies of T cells producing interferon gamma (IFN-γ) are presented. Statistical significance is indicated as follows: *P < 0.05, **P < 0.01.

## Notes

### Clinical Trial

NCT04871737

### Author Declarations

The study was approved by the Federal Commission for the Protection against Sanitary Risks (COFEPRIS) in Mexico under number 213300410A0063/2021, after approval by the Ethics, Biosafety and Research Committees of the clinical research site Hospital Medica Sur (03-2021-CI/CEI/CB-156) in full compliance of the Mexican regulation and under the principles of the Declaration of Helsinki and Good Clinical Practice.

## References

1. Zhou P, Yang XL, Wang XG, Hu B, Zhang L, Zhang W, Si HR, Zhu Y, Li B, Huang CL, Chen HD, Chen J, Luo Y, Guo H, Jiang RD, Liu MQ, Chen Y, Shen XR, Wang X, Zheng XS, Zhao K, Chen QJ, Deng F, Liu LL, Yan B, Zhan FX, Wang YY, Xiao GF, Shi ZL. 2020. A pneumonia outbreak associated with a new coronavirus of probable bat origin. Nature.

2. Zhu N, Zhang D, Wang W, Li X, Yang B, Song J, Zhao X, Huang B, Shi W, Lu R, Niu P, Zhan F, Ma X, Wang D, Xu W, Wu G, Gao GF, Tan W, Team CNCIaR. 2020. A Novel Coronavirus from Patients with Pneumonia in China, 2019. N Engl J Med 382:727–733.

3. Krammer F. 2020. SARS-CoV-2 vaccines in development. Nature 586:516–527.

4. Su F, Patel GB, Hu S, Chen W. 2016. Induction of mucosal immunity through systemic immunization: Phantom or reality? Hum Vaccin Immunother 12:1070–9.

5. Liu Y, Rocklöv J. 2021. The reproductive number of the Delta variant of SARS-CoV-2 is far higher compared to the ancestral SARS-CoV-2 virus. J Travel Med 28.

6. Andrews N, Stowe J, Kirsebom F, Toffa S, Rickeard T, Gallagher E, Gower C, Kall M, Groves N, O’Connell A-M, Simons D, Blomquist PB, Zaidi A, Nash S, Aziz NIBA, Thelwall S, Dabrera G, Myers R, Amirthalingam G, Gharbia S, Barrett JC, Elson R, Ladhani SN, Ferguson N, Zambon M, Campbell CN, Brown K, Hopkins S, Chand M, Ramsay M, Bernal JL. 2021. Effectiveness of COVID-19 vaccines against the Omicron (B.1.1.529) variant of concern. medRxiv:2021.12.14.21267615.

7. Carreño JM, Alshammary H, Tcheou J, Singh G, Raskin A, Kawabata H, Sominsky L, Clark J, Adelsberg DC, Bielak D, Gonzalez-Reiche AS, Dambrauskas N, Vigdorovich V, Srivastava K, Sather DN, Sordillo EM, Bajic G, van Bakel H, Simon V, Krammer F, Group PPS. 2021. Activity of convalescent and vaccine serum against SARS-CoV-2 Omicron. Nature.

8. Pulliam JRC, van Schalkwyk C, Govender N, von Gottberg A, Cohen C, Groome MJ, Dushoff J, Mlisana K, Moultrie H. 2021. Increased risk of SARS-CoV-2 reinfection associated with emergence of the Omicron variant in South Africa. medRxiv:2021.11.11.21266068.

9. Brandal LT, MacDonald E, Veneti L, Ravlo T, Lange H, Naseer U, Feruglio S, Bragstad K, Hungnes O, Ødeskaug LE, Hagen F, Hanch-Hansen KE, Lind A, Watle SV, Taxt AM, Johansen M, Vold L, Aavitsland P, Nygård K, Madslien EH. 2021. Outbreak caused by the SARS-CoV-2 Omicron variant in Norway, November to December 2021. Eurosurveillance 26:2101147.

10. Gu H, Krishnan P, Ng DYM, Chang LDJ, Liu GYZ, Cheng SSM, Hui MMY, Fan MCY, Wan JHL, Lau LHK, Cowling BJ, Peiris M, Poon LLM. 2021. Probable Transmission of SARS-CoV-2 Omicron Variant in Quarantine Hotel, Hong Kong, China, November 2021. Emerg Infect Dis 28.

11. Wilhelm A, Widera M, Grikscheit K, Toptan T, Schenk B, Pallas C, Metzler M, Kohmer N, Hoehl S, Helfritz FA, Wolf T, Goetsch U, Ciesek S. 2021. Reduced Neutralization of SARS-CoV-2 Omicron Variant by Vaccine Sera and Monoclonal Antibodies. medRxiv:2021.12.07.21267432.

12. Rössler A, Riepler L, Bante D, Laer Dv, Kimpel J. 2021. SARS-CoV-2 B.1.1.529 variant (Omicron) evades neutralization by sera from vaccinated and convalescent individuals. medRxiv:2021.12.08.21267491.

13. Cele S, Jackson L, Khan K, Khoury DS, Moyo-Gwete T, Tegally H, Scheepers C, Amoako D, Karim F, Bernstein M, Lustig G, Archary D, Smith M, Ganga Y, Jule Z, Reedoy K, San JE, Hwa S-H, Giandhari J, Blackburn J, Gosnell BI, Karim SA, Hanekom W, NGS-SA, Team C-K, von Gottberg A, Bhiman J, Lessells R, Moosa M-YS, Davenport MP, de Oliveira T, Moore PL, Sigal A. 2021. SARS-CoV-2 Omicron has extensive but incomplete escape of Pfizer BNT162b2 elicited neutralization and requires ACE2 for infection. medRxiv:2021.12.08.21267417.

14. Rosenberg ES, Holtgrave DR, Dorabawila V, Conroy M, Greene D, Lutterloh E, Backenson B, Hoefer D, Morne J, Bauer U, Zucker HA. 2021. New COVID-19 Cases and Hospitalizations Among Adults, by Vaccination Status - New York, May 3-July 25, 2021. MMWR Morb Mortal Wkly Rep 70:1306–1311.

15. Fowlkes A, Gaglani M, Groover K, Thiese MS, Tyner H, Ellingson K, Cohorts H-R. 2021. Effectiveness of COVID-19 Vaccines in Preventing SARS-CoV-2 Infection Among Frontline Workers Before and During B.1.617.2 (Delta) Variant Predominance - Eight U.S. Locations, December 2020-August 2021. MMWR Morb Mortal Wkly Rep 70:1167–1169.

16. Csatary LK, Eckhardt S, Bukosza I, Czegledi F, Fenyvesi C, Gergely P, Bodey B, Csatary CM. 1993. Attenuated veterinary virus vaccine for the treatment of cancer. Cancer Detect Prev 17:619–27.

17. Wagner S, Csatary CM, Gosztonyi G, Koch HC, Hartmann C, Peters O, Hernáiz-Driever P, Théallier-Janko A, Zintl F, Längler A, Wolff JE, Csatary LK. 2006. Combined treatment of pediatric high-grade glioma with the oncolytic viral strain MTH-68/H and oral valproic acid. APMIS 114:731–43.

18. Shirvani E, Samal SK. 2020. Newcastle Disease Virus as a Vaccine Vector for SARS-CoV-2. Pathogens 9.

19. Park MS, García-Sastre A, Cros JF, Basler CF, Palese P. 2003. Newcastle disease virus V protein is a determinant of host range restriction. J Virol 77:9522–32.

20. Kong D, Wen Z, Su H, Ge J, Chen W, Wang X, Wu C, Yang C, Chen H, Bu Z. 2012. Newcastle disease virus-vectored Nipah encephalitis vaccines induce B and T cell responses in mice and long-lasting neutralizing antibodies in pigs. Virology 432:327–35.

21. Martinez-Sobrido L, Gitiban N, Fernandez-Sesma A, Cros J, Mertz SE, Jewell NA, Hammond S, Flano E, Durbin RK, García-Sastre A, Durbin JE. 2006. Protection against respiratory syncytial virus by a recombinant Newcastle disease virus vector. J Virol 80:1130–9.

22. Nakaya T, Cros J, Park MS, Nakaya Y, Zheng H, Sagrera A, Villar E, Garcia-Sastre A, Palese P. 2001. Recombinant Newcastle disease virus as a vaccine vector. J Virol 75:11868–73.

23. Bukreyev A, Huang Z, Yang L, Elankumaran S, St Claire M, Murphy BR, Samal SK, Collins PL. 2005. Recombinant newcastle disease virus expressing a foreign viral antigen is attenuated and highly immunogenic in primates. J Virol 79:13275–84.

24. Kortekaas J, de Boer SM, Kant J, Vloet RP, Antonis AF, Moormann RJ. 2010. Rift Valley fever virus immunity provided by a paramyxovirus vaccine vector. Vaccine 28:4394–401.

25. Sun W, McCroskery S, Liu WC, Leist SR, Liu Y, Albrecht RA, Slamanig S, Oliva J, Amanat F, Schäfer A, Dinnon KH, Innis BL, García-Sastre A, Krammer F, Baric RS, Palese P. 2020. A Newcastle Disease Virus (NDV) Expressing a Membrane-Anchored Spike as a Cost-Effective Inactivated SARS-CoV-2 Vaccine. Vaccines (Basel) 8.

26. Sun W, Leist SR, McCroskery S, Liu Y, Slamanig S, Oliva J, Amanat F, Schäfer A, Dinnon KH, García-Sastre A, Krammer F, Baric RS, Palese P. 2020. Newcastle disease virus (NDV) expressing the spike protein of SARS-CoV-2 as a live virus vaccine candidate. EBioMedicine 62:103132.

27. Sun W, Liu Y, Amanat F, González-Domínguez I, McCroskery S, Slamanig S, Coughlan L, Rosado V, Lemus N, Jangra S, Rathnasinghe R, Schotsaert M, Martinez J, Sano K, Mena I, Innis BL, Wirachwong P, Thai DH, Oliveira RDN, Scharf R, Hjorth R, Raghunandan R, Krammer F, García-Sastre A, Palese P. 2021. A Newcastle disease virus-vector expressing a prefusion-stabilized spike protein of SARS-CoV-2 induces protective immune responses against prototype virus and variants of concern in mice and hamsters. bioRxiv:2021.07.06.451301.

28. Sun W, Liu Y, Amanat F, González-Domínguez I, McCroskery S, Slamanig S, Coughlan L, Rosado V, Lemus N, Jangra S, Rathnasinghe R, Schotsaert M, Martinez JL, Sano K, Mena I, Innis BL, Wirachwong P, Thai DH, Oliveira RDN, Scharf R, Hjorth R, Raghunandan R, Krammer F, García-Sastre A, Palese P. 2021. A Newcastle disease virus expressing a stabilized spike protein of SARS-CoV-2 induces protective immune responses. Nat Commun 12:6197.

29. Hsieh CL, Goldsmith JA, Schaub JM, DiVenere AM, Kuo HC, Javanmardi K, Le KC, Wrapp D, Lee AG, Liu Y, Chou CW, Byrne PO, Hjorth CK, Johnson NV, Ludes-Meyers J, Nguyen AW, Park J, Wang N, Amengor D, Lavinder JJ, Ippolito GC, Maynard JA, Finkelstein IJ, McLellan JS. 2020. Structure-based design of prefusion-stabilized SARS-CoV-2 spikes. Science 369:1501–1505.

30. Sparrow E, Wood JG, Chadwick C, Newall AT, Torvaldsen S, Moen A, Torelli G. 2021. Global production capacity of seasonal and pandemic influenza vaccines in 2019. Vaccine 39:512–520.

31. Lara-Puente JH, Carreño JM, Sun W, Suárez-Martínez A, Ramírez-Martínez L, Quezada-Monroy F, Paz-De la Rosa G, Vigueras-Moreno R, Singh G, Rojas-Martínez O, Chagoya-Cortés HE, Sarfati-Mizrahi D, Soto-Priante E, López-Macías C, Krammer F, Castro-Peralta F, Palese P, García-Sastre A, Lozano-Dubernard B. 2021. Safety and Immunogenicity of a Newcastle Disease Virus Vector-Based SARS-CoV-2 Vaccine Candidate, AVX/COVID-12-HEXAPRO (Patria), in Pigs. mBio:e0190821.

32. Pitisuttithum P, Luvira V, Lawpoolsri S, Muangnoicharoen S, Kamolratanakul S, Sivakorn C, Narakorn P, Surichan S, Prangpratanporn S, Puksuriwong S, Lamola S, Mercer LD, Raghunandan R, Sun W, Liu Y, Carreño JM, Scharf R, Phumratanaprapin W, Amanat F, Gagnon L, Hsieh CL, Kaweepornpoj R, Khan S, Lal M, McCroskery S, McLellan J, Mena I, Meseck M, Phonrat B, Sabmee Y, Singchareon R, Slamanig S, Suthepakul N, Tcheou J, Thantamnu N, Theerasurakarn S, Tran S, Vilasmongkolchai T, White JA, Garcia-Sastre A, Palese P, Krammer F, Poopipatpol K, Wirachwong P, Hjorth R, Innis BL. 2021. Safety and Immunogenicity of an Inactivated Recombinant Newcastle Disease Virus Vaccine Expressing SARS-CoV-2 Spike: Interim Results of a Randomised, Placebo-Controlled, Phase 1/2 Trial. medRxiv.

33. Tcheou J, Raskin A, Singh G, Kawabata H, Bielak D, Sun W, González-Domínguez I, Sather DN, García-Sastre A, Palese P, Krammer F, Carreño JM. 2021. Safety and Immunogenicity Analysis of a Newcastle Disease Virus (NDV-HXP-S) Expressing the Spike Protein of SARS-CoV-2 in Sprague Dawley Rats. Front Immunol 12:791764.

34. Cedro-Tanda A, Gómez-Romero L, Alcaraz N, de Anda-Jauregui G, Peñaloza F, Moreno B, Escobar-Arrazola MA, Ramirez-Vega OA, Munguia-Garza P, Garcia-Cardenas F, Cisneros-Villanueva M, Moreno-Camacho JL, Rodriguez-Gallegos J, Luna-Ruiz Esparza MA, Fernández Rojas MA, Mendoza-Vargas A, Reyes-Grajeda JP, Campos-Romero A, Angulo O, Ruiz R, Sheinbaum-Pardo C, Sifuentes-Osornio J, Kershenobich D, Hidalgo-Miranda A, Herrera LA. 2021. The Evolutionary Landscape of SARS-CoV-2 Variant B.1.1.519 and Its Clinical Impact in Mexico City. Viruses 13.

35. Harritshøj LH, Gybel-Brask M, Afzal S, Kamstrup PR, Jørgensen CS, Thomsen MK, Hilsted L, Friis-Hansen L, Szecsi PB, Pedersen L, Nielsen L, Hansen CB, Garred P, Korsholm TL, Mikkelsen S, Nielsen KO, Møller BK, Hansen AT, Iversen KK, Nielsen PB, Hasselbalch RB, Fogh K, Norsk JB, Kristensen JH, Schønning K, Kirkby NS, Nielsen ACY, Landsy LH, Loftager M, Holm DK, Nilsson AC, Sækmose SG, Grum-Schwensen B, Aagaard B, Jensen TG, Nielsen DM, Ullum H, Dessau RB. 2021. Comparison of 16 Serological SARS-CoV-2 Immunoassays in 16 Clinical Laboratories. J Clin Microbiol 59.

36. Tan CW, Chia WN, Qin X, Liu P, Chen MI, Tiu C, Hu Z, Chen VC, Young BE, Sia WR, Tan YJ, Foo R, Yi Y, Lye DC, Anderson DE, Wang LF. 2020. A SARS-CoV-2 surrogate virus neutralization test based on antibody-mediated blockage of ACE2-spike protein-protein interaction. Nat Biotechnol 38:1073–1078.

37. O’Hara DM, Xu Y, Liang Z, Reddy MP, Wu DY, Litwin V. 2011. Recommendations for the validation of flow cytometric testing during drug development: II assays. J Immunol Methods 363:120–34.

38. Earle KA, Ambrosino DM, Fiore-Gartland A, Goldblatt D, Gilbert PB, Siber GR, Dull P, Plotkin SA. 2021. Evidence for antibody as a protective correlate for COVID-19 vaccines. Vaccine.

39. Gilbert PB, Montefiori DC, McDermott A, Fong Y, Benkeser D, Deng W, Zhou H, Houchens CR, Martins K, Jayashankar L, Castellino F, Flach B, Lin BC, O’Connell S, McDanal C, Eaton A, Sarzotti-Kelsoe M, Lu Y, Yu C, Borate B, van der Laan LWP, Hejazi N, Huynh C, Miller J, El Sahly HM, Baden LR, Baron M, De La Cruz L, Gay C, Kalams S, Kelley CF, Kutner M, Andrasik MP, Kublin JG, Corey L, Neuzil KM, Carpp LN, Pajon R, Follmann D, Donis RO, Koup RA, Assays obotI, Moderna I, Efficacy CVPNC, Teams aUSGCB. 2021. Immune Correlates Analysis of the mRNA-1273 COVID-19 Vaccine Efficacy Trial. medRxiv:2021.08.09.21261290.

40. McMahan K, Yu J, Mercado NB, Loos C, Tostanoski LH, Chandrashekar A, Liu J, Peter L, Atyeo C, Zhu A, Bondzie EA, Dagotto G, Gebre MS, Jacob-Dolan C, Li Z, Nampanya F, Patel S, Pessaint L, Van Ry A, Blade K, Yalley-Ogunro J, Cabus M, Brown R, Cook A, Teow E, Andersen H, Lewis MG, Lauffenburger DA, Alter G, Barouch DH. 2021. Correlates of protection against SARS-CoV-2 in rhesus macaques. Nature 590:630–634.

41. Pitisuttithum P, Luvira V, Lawpoolsri S, Muangnoicharoen S, Kamolratanakul S, Sivakorn C, Narakorn P, Surichan S, Prangpratanporn S, Puksuriwong S, Lamola S, Mercer LD, Raghunandan R, Sun W, Liu Y, Carreño JM, Scharf R, Phumratanaprapin W, Amanat F, Gagnon L, Hsieh C-L, Kaweepornpoj R, Khan S, Lal M, McCroskery S, McLellan J, Mena I, Meseck M, Phonrat B, Sabmee Y, Singchareon R, Slamanig S, Suthepakul N, Tcheou J, Thantamnu N, Theerasurakarn S, Tran S, Vilasmongkolchai T, White JA, Garcia-Sastre A, Palese P, Krammer F, Poopipatpol K, Wirachwong P, Hjorth R, Innis BL. 2021. Safety and Immunogenicity of an Inactivated Recombinant Newcastle Disease Virus Vaccine Expressing SARS-CoV-2 Spike: Interim Results of a Randomised, Placebo-Controlled, Phase 1/2 Trial. medRxiv:2021.09.17.21263758.

42. Jackson LA, Anderson EJ, Rouphael NG, Roberts PC, Makhene M, Coler RN, McCullough MP, Chappell JD, Denison MR, Stevens LJ, Pruijssers AJ, McDermott A, Flach B, Doria-Rose NA, Corbett KS, Morabito KM, O’Dell S, Schmidt SD, Swanson PA, Padilla M, Mascola JR, Neuzil KM, Bennett H, Sun W, Peters E, Makowski M, Albert J, Cross K, Buchanan W, Pikaart-Tautges R, Ledgerwood JE, Graham BS, Beigel JH, Group m-S. 2020. An mRNA Vaccine against SARS-CoV-2 - Preliminary Report. N Engl J Med.

43. Polack FP, Thomas SJ, Kitchin N, Absalon J, Gurtman A, Lockhart S, Perez JL, Pérez Marc G, Moreira ED, Zerbini C, Bailey R, Swanson KA, Roychoudhury S, Koury K, Li P, Kalina WV, Cooper D, Frenck RW, Hammitt LL, Türeci Ö, Nell H, Schaefer A, Ünal S, Tresnan DB, Mather S, Dormitzer PR, Şahin U, Jansen KU, Gruber WC, Group CCT. 2020. Safety and Efficacy of the BNT162b2 mRNA Covid-19 Vaccine. N Engl J Med.

44. Voysey M, Clemens SAC, Madhi SA, Weckx LY, Folegatti PM, Aley PK, Angus B, Baillie VL, Barnabas SL, Bhorat QE, Bibi S, Briner C, Cicconi P, Collins AM, Colin-Jones R, Cutland CL, Darton TC, Dheda K, Duncan CJA, Emary KRW, Ewer KJ, Fairlie L, Faust SN, Feng S, Ferreira DM, Finn A, Goodman AL, Green CM, Green CA, Heath PT, Hill C, Hill H, Hirsch I, Hodgson SHC, Izu A, Jackson S, Jenkin D, Joe CCD, Kerridge S, Koen A, Kwatra G, Lazarus R, Lawrie AM, Lelliott A, Libri V, Lillie PJ, Mallory R, Mendes AVA, Milan EP, Minassian AM, et al. 2020. Safety and efficacy of the ChAdOx1 nCoV-19 vaccine (AZD1222) against SARS-CoV-2: an interim analysis of four randomised controlled trials in Brazil, South Africa, and the UK. Lancet.

45. Folegatti PM, Ewer KJ, Aley PK, Angus B, Becker S, Belij-Rammerstorfer S, Bellamy D, Bibi S, Bittaye M, Clutterbuck EA, Dold C, Faust SN, Finn A, Flaxman AL, Hallis B, Heath P, Jenkin D, Lazarus R, Makinson R, Minassian AM, Pollock KM, Ramasamy M, Robinson H, Snape M, Tarrant R, Voysey M, Green C, Douglas AD, Hill AVS, Lambe T, Gilbert SC, Pollard AJ, Group OCVT. 2020. Safety and immunogenicity of the ChAdOx1 nCoV-19 vaccine against SARS-CoV-2: a preliminary report of a phase 1/2, single-blind, randomised controlled trial. Lancet 396:467–478.

